# Covid-19: Qualitative Change in the Behavior of the “Virus vs Human” System - From Limit Cycle to Sustained Focus

**DOI:** 10.1101/2022.09.28.22280472

**Authors:** Alexander V. Sokolov

**Affiliations:** Institute for information transmission problem (Kharkevitch Insitute) RAS, Bolshoy Karetny per. 19, build.1, Moscow 127051 Russia; Vernadsky Institute of Geochemistry and Analytical Chemistry of RAS, Kosygin street 19, Moscow 119991 Russia

**Keywords:** incidence, COVID-19, immunity, modeling, bifurcation, balanced identification

## Abstract

The high contagiousness of the latest strains of Covid-19 qualitatively changes the behavior of the “virus vs human” system. Numerical experiments with a model of the Covid-19 epidemic in Moscow have shown that a reproduction number R0 of about 4 is critical, defining a qualitative change in the dynamics of the epidemic. Below this value (observed until 2022), the long-term forecast tends to undamped oscillations; above this value, it is described by damped oscillations: amplitudes of the epidemic waves get smaller and smaller, with a constant, very high background level of morbidity (or high-intensity vaccination) that maintains the state of natural immunity at a level close to 100% (reaching 93.7% for the current R0 value of about 16). At the limit, the system tends to a stable equilibrium point. Here we consider a reduced model of epidemic dynamics. Its study (search for equilibrium solutions, analysis of their stability, construction a bifurcation diagram and a phase portrait) confirms the presence of points of qualitative change in the behavior of the “virus vs human” system (bifurcation points). Some practical results for Moscow are given. A further increase in the contagiousness of the virus does not change the picture significantly, thus more infectious strains are not to be feared. The key parameter of the study is the function of the immunity level depending on the time after the disease. The damping of omicron waves (oscillations), observed recently in many countries, is a confirmation of the correctness of the accepted hypotheses.

## Introduction

Figure 1 shows the results of forecasts of new detected cases of infection (nC), obtained on the basis of a model, identified on real data for Moscow [1, 2], dated 04/27/2022 and 09/06/2022. In the first figure (1a), the dynamics tends to undamped oscillations with a period of about 8 months and with a maximum of about 15 thousand people, in the second (1b), it enters the damped oscillations mode around the (background) value of 3.5 thousand people and with a period of about 7 months, with the amplitude of each wave 30% lower than the previous one.

**Figure 1.**
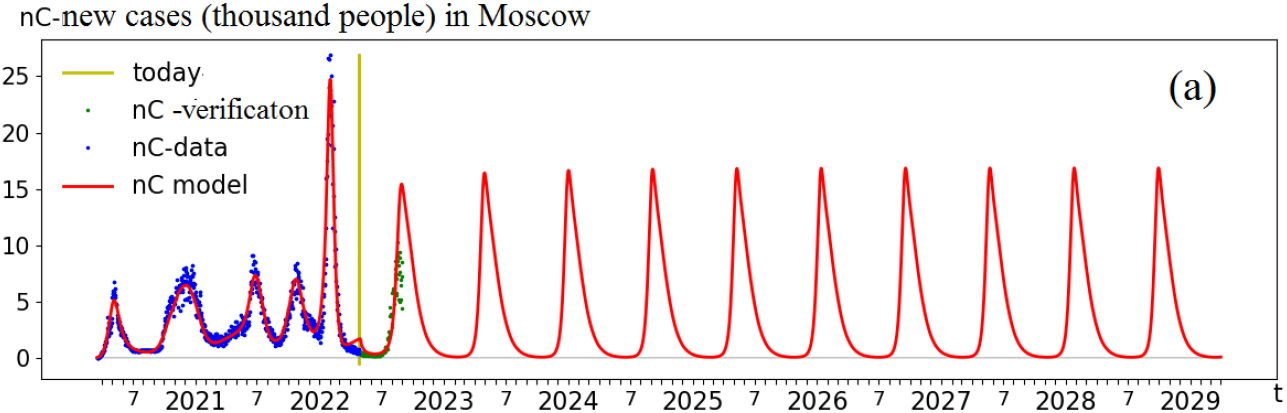
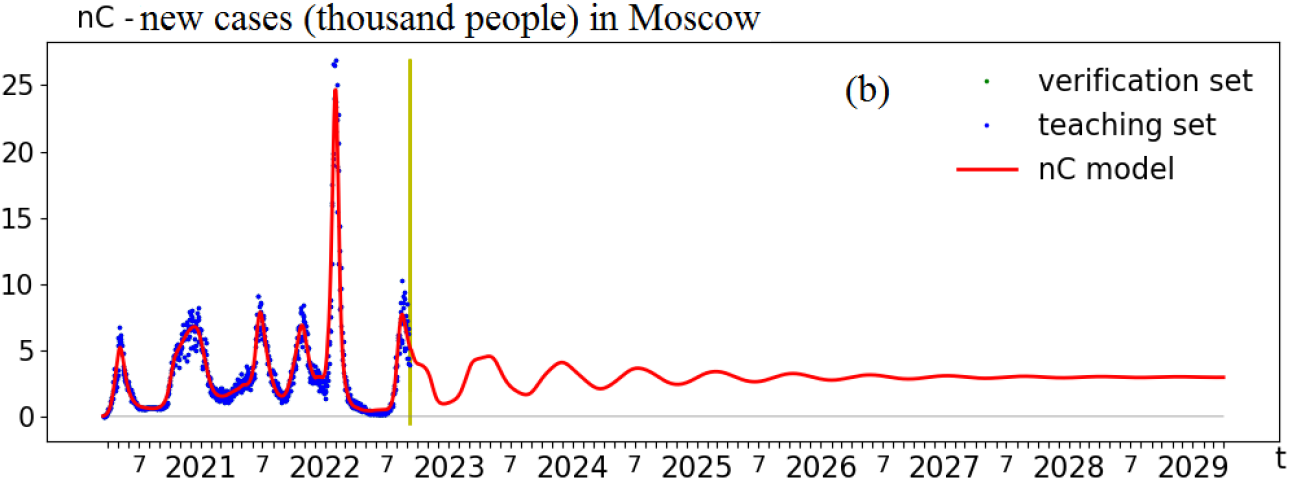
Long-term forecasts for Moscow: (a) Forecast from 04/27/2022 with vaccination rate of 20000/day, testing rate of 1.4 tests/1000/day, and reproduction number R0=3.8. (b) Forecast from 09/06/2022 with vaccination rate of 2000/day, testing rate of 2 tests/1000/day, and reproduction number of R0=16.6. Start of simulation: 19/03/2020. Marks on the t-axis correspond to months and years. The vertical yellow line shows the start of the forecast. The model curves (shown in red) obtained (or identified) on the training data set (blue dots) can be compared with the testing set (green dots).

The main parameter determining the qualitative differences in the dynamics of forecasts is the reproduction number R0. In the first forecast, the value of R0=3.8 (or 2.7, taking into account vaccination) in the second – R0=16.6 (15.6, taking into account vaccination).

Thus, a change in the contagiousness of the virus leads to a qualitative change in the behavior of the “virus vs human” system.

The examples above emphasize the importance of a more complex study of the model used: the determination of stationary states and limit cycles, their stability depending on the critical (bifurcation) parameter R0, i.e. construction of the bifurcation diagram of the system.

### Reduced model and its study

Under certain, fairly simple assumptions, such as the absence of patient’s isolation, zero migration, etc., the model used [1, 2] can be significantly simplified and reduced to the form:

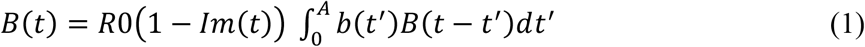

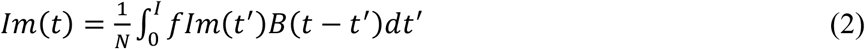

where *B(t)* is the number of infected people at time *t, R0* – the reproduction number, *Im(t)* – the proportion of the population with immunity, *A* – the maximum duration of the disease (14 days), b(*t*′) – the normalized 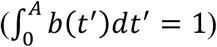 infectivity as a function of the duration of the disease *t*′ (number of days since the infection), *N* – the population size, *I* – the maximum duration of immunity, *fIm*(*t*′) – the proportion of infected people who retained immunity through *t*′ days since the infection.

Equation (1) describes the number of the new infected people as the product of the reproductive number *R0* (how many are infected by one infected person) by the proportion of the population without immunity *(1-Im(t))* and by the number of infected (with different times from the onset of the disease), summed up taking into account the difference in infectivity, depending on the time from the onset of the disease.

Equation (2) describes the proportion of natural immunity carriers as the number of infected people who retained immunity at the moment *t*, divided by the total population *N*.

The functions *b(t)* and *fIm(t)* are shown in Figure 2. They were obtained earlier when identifying the full model [1, 2] for Moscow. It is these functions, together with the parameter R0, that determine the behavior of the system.

**Figure 2.**
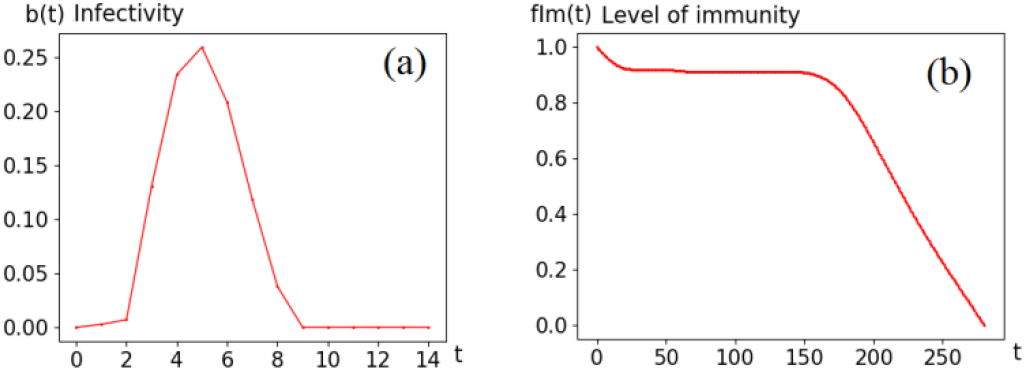
The main functions that determine the dynamics of the model (1)-(2): (a) Normalized (the integral being 1) infectivity as a function of the duration of infection (days). (b) The level of immunity (1 corresponds to 100%) as a function of the time since the onset of the disease (days).

Let us briefly present the results of a mathematical study of system (1)-(2). It is easy to find stationary (equilibrium) solutions:

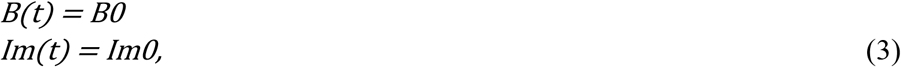

with is the equilibrium point:

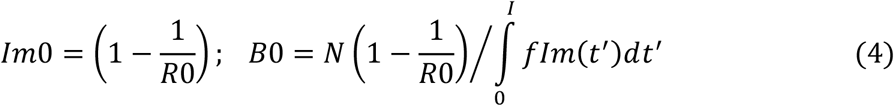

Put (2) into (1) to study stability,

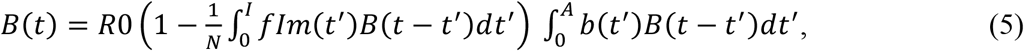

after the equilibrium point region linearization (5): B(t) = B0 + δ (t) and, neglecting the terms of the 2nd order, the linear integral equation is obtained

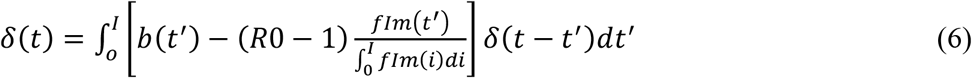

or

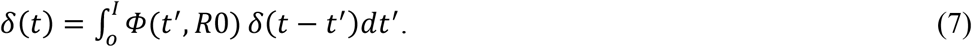

Substituting *δ*(*t*) = *e*^*λ t*^:

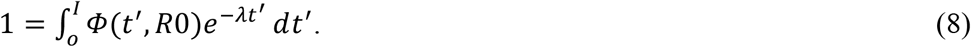

If for all solutions λ of equation (8) *Re(λ)* < 0, then the equilibrium (4) of the system of equations (1)-(2) is stable. Otherwise, if there exists λ with *Re(λ)* > 0, the equilibrium is unstable. Thus, one can determine the stability of stationary solutions for various *R0*.

The considered equations do not have analytical solutions (especially since the functions *b(t)* and *fIm(t)* are the result of another study and are given numerically), therefore, numerical models approximating them, which are not given here, were studied. Let us present some of the results obtained on the basis of the balanced identification technology [3]

### Numerical results and discussion

*Note. The numbers below are approximate and may be adjusted as needed*.

Figure 3 shows the bifurcation diagram of the studied system (1)-(2).

**Figure 3.**
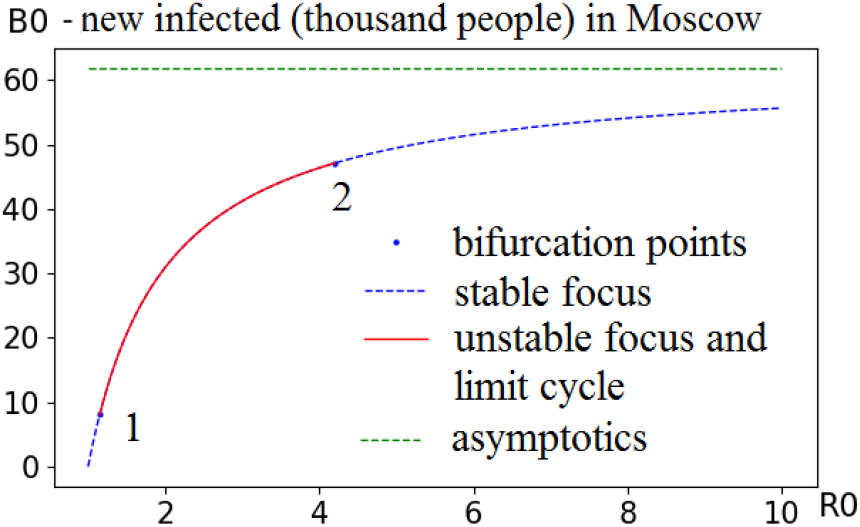
Bifurcation diagram.

With an increase in R0, at point 1, a stable node is transformed into an unstable node and a cycle. At point 2, the reverse occurs – the unstable node and the cycle turn into a stable node.

Similar constructions look more clearly in the phase space *B*× *Im*.

Figure 4 shows the phase portrait of system (1)-(2). The point on the far left of the line with coordinates (0,0) corresponds to *R0* ≤ 1. Up to the bifurcation point 1 (*R0* value about 1.16) the line corresponds to a stable focus. The example trajectory shown here (blue stable focus) corresponds to *R0*=1.1. At bifurcation point 1, a qualitative change in the dynamic properties of the system occurs: the focus becomes unstable and a limit cycle appears. The given example of the trajectory corresponds to *R0*=1.3. As we move to point 2, the amplitude of the cycle first increases (for example, at *R0*=3 the *B* amplitude reaches 700 thousand), and then it starts to fall (at *R0*=3.6 the *B* amplitude is about 80 thousand) and the cycle becomes more complex (with self-intersections), and closer to point 2, the motion of the system possibly becomes chaotic (this statement requires additional research). After the bifurcation point 2, the qualitative behavior of the system is again described by a stable focus. The given example corresponds to *R0*=16.

**Figure 4.**
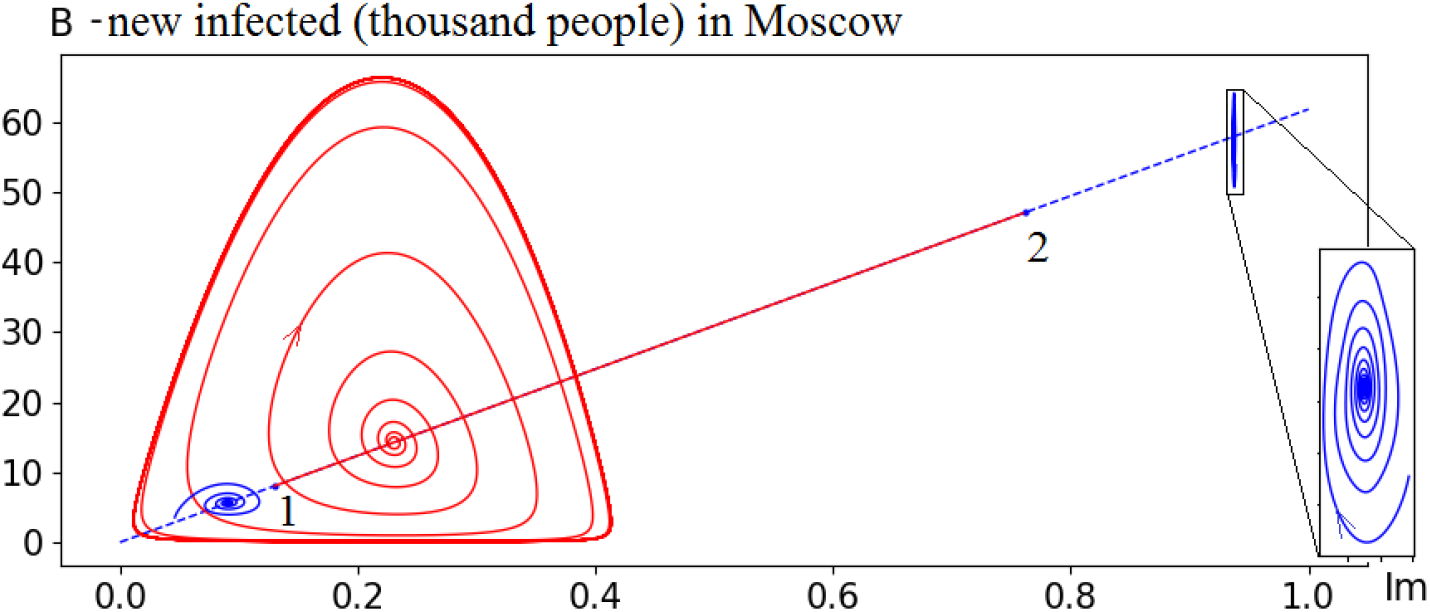
Phase portrait of the system (1)-(2): 1 and 2 are bifurcation points, stable foci are blue, unstable focus and limit cycle are red.

#### Some practical results

In the limit, the system tends to a stable equilibrium point. The level of immunity at this point is determined by the formula (R0-1) / R0 * 100%. In order to stop the spread of a virus with a (basic) reproduction number R0=16, it is necessary to reduce it to 1, i.e. 15 people (out of 16) should be immune and therefore (R0-1)/R0.

The table shows stationary solutions (equilibrium points) for Moscow, calculated for various values of R0.

**Table.** Equilibrium points of system (1)-(2) for Moscow.

| $R_0$ | $B_0$ (thousand people per day) | $Im_0$ (%) |
| --- | --- | --- |
| 5 | 49,6 | 80 |
| 10 | 55,8 | 90 |
| 15 | 57,9 | 93,3 |
| 20 | 58,9 | 95 |

The analysis of the table shows that with values of the reproduction number of more than 10, stationary solutions differ slightly from each other, with more than 90% of carriers of natural immunity and the total number of cases (detected and undetected summed) from 55 to 60 thousand people per day. Thus, according to the model, more infectious viruses are not to be feared.

With the current percentage of asymptomatic infected, the level of testing, the level of treatment and self-treatment, etc. for 60 thousand cases per day, rough estimates of other indicators are:

- detected cases: about 3 thousand people per day,
- hospitalized: about 150 people per day,
- deaths: about 20 people per day.

## Conclusion

A mathematical study of the reduced model (1)-(2) of the Covid-19 epidemic in Moscow and the corresponding numerical calculations confirmed the assumption of a qualitative change in the behavior of the “virus vs human” system with an increase in the virus infectiousness. The critical value of the reproduction number *R0* is about 4. Below this value (which we observed until 2022), the long-term forecast tends to undamped oscillations (limiting cycle), above this value, it is described by damped oscillations (stable focus): the amplitudes of epidemic waves lessen with a constant, very high background level of morbidity, maintaining the state of natural immunity at a level close to 100%. For the current situation in Moscow (September 2022), the period of waves is about 7.5 months, the amplitude of each wave is 30% lower than the previous one.

The main assumption of this study is the constancy of the function of immunity level (see Fig. 2b), different strains included. The results presented here and more detailed studies of strain competition [4] allow us to consider this assumption justified for the time being. The common assumption that each new variant (strain) of the virus raises a wave of morbidity is not always the true – “stealth”, which appeared towards the end of the winter wave of 2022, managed to displace the initial version of “omicron”, but could not reproduce fast enough due to the high proportion of the immunized. Another example, the “BA4/5” strain appeared in May-June 2022, but entered the rapid growth stage only in July, with immunity from the winter wave subsiding.

Within the framework of the considered models, the statement about the future seasonal nature of the Covid-19 epidemic seems doubtful. If seasonality means a seasonal increase in the incidence due to the autumn increase in contacts between people, then with the current high level of infectiousness and a high proportion of the immunized population, an increase in contacts by, say, 30% (compared to summer) will only lead to a small wave, with an increase of the number of identified infected by about 10%. Real statistical data also shows no seasonality. For example, the last wave of the virus in Moscow occurred in August 2022.

There is hope that our immunity will learn to “remember” the virus for longer (now, according to calculations [1,2] it is about 200 days), and then the “tribute” the mankind pays for maintaining immunity will become significantly less than the above figures. Another hope is vaccination, including more effective vaccines.

## Data Availability

All data produced in the present work are contained in the manuscript

## Acknowledgments

This research was carried out with the financial support of the Russian Scientific Foundation within the framework of the scientific project (grant) 22-11-00317. This work has been carried out using computing resources of the federal collective usage center Complex for Simulation and Data Processing for Megascience Facilities at NRC “Kurchatov Institute”.

